# Sex-Specific Hemostatic Responses and Diagnostic Potential of Platelet Distribution Width (PDW) and D-Dimer in Mild COVID-19, Malaria, and Co-Infection in a Tropical Setting: A Case-Control Study in Port Harcourt, Nigeria

**DOI:** 10.64898/2026.06.21.26356162

**Authors:** E. S. Ekprikpo, Z. A. Jeremiah, S. U. Ken-Ezihuo

## Abstract

**Background:** In malaria-endemic tropical regions, the overlapping coagulopathy in COVID-19 and malaria poses diagnostic and prognostic challenges, particularly with potential sex differences. This study evaluated sex-specific variations in platelet indices and fibrinolytic markers and assessed the utility of Platelet Distribution Width (PDW) and D-dimer in mild/asymptomatic cases.

**Methods:** A case-control study was conducted with 220 participants (55 each in healthy controls, malaria-positive, COVID-19-positive, and COVID-19+malaria co-infected groups), aged 20–65 years, in Port Harcourt, Nigeria. Platelet indices were analysed using Sysmex XP-300 haematology analyser, while D-dimer and fibrinogen were measured by ELISA. Data were analysed using SAS 9.4 with ANOVA, Tukey’s HSD, Pearson correlation, and sex-stratified comparisons.

**Results:** PDW was significantly elevated in all infected groups compared to controls (malaria: 15.21 ± 0.22 fL; COVID-19: 15.21 ± 0.22 fL; co-infection: 15.61 ± 0.21 fL vs. control: 13.26 ± 0.17 fL; F=25.850, p < 0.001). D-dimer levels were highest in the co-infected group (553.42 ± 59.74 ng/ml, F=2.816, p = 0.040). No significant changes were observed in other platelet indices or fibrinogen across groups. No significant correlation existed between platelet indices and the fibrinolytic markers. Males exhibited significantly higher D-dimer levels across all infected groups (p < 0.05) and higher fibrinogen in COVID-19 subjects (p = 0.036). Sex exerted a stronger influence on parameters than age.

**Conclusion:** Males show heightened fibrinolytic activation in COVID-19 and malaria co-infection. PDW and D-dimer are promising, cost-effective biomarkers for screening mild infections in resource-limited tropical settings.

## Introduction

The coexistence of COVID-19 and malaria in tropical regions creates a syndemic with overlapping clinical features and shared pathways of coagulopathy (Gutman *et al*., 2020). Both conditions are associated with endothelial dysfunction, platelet activation, and fibrinolytic imbalance, increasing the risk of thrombosis (Li *et al*., 2024). While previous analyses from this cohort described general platelet derangements and exacerbated fibrinolytic responses in co-infection, sex-specific patterns and the practical diagnostic value of routine haematological parameters remain underexplored (Bohn *et al*., 2020).

This study builds on the original dataset to provide a focused examination of sex-specific hemostatic responses and the potential utility of Platelet Distribution Width (PDW) a simple, inexpensive marker of platelet anisocytosis alongside D-dimer for early detection and risk stratification in mild/asymptomatic cases in a high-burden tropical setting.

## Methods

### Study Design and Participants

This comparative case-control study utilised data from 220 adults (55 per group) recruited in Port Harcourt, Rivers State, Nigeria. Groups included: apparently healthy controls, malaria-positive, COVID-19-positive, and COVID-19 + malaria co-infected subjects. Inclusion criteria were age 20–65 years and confirmed infection status. Ethical clearance was obtained from the Rivers State Ministry of Health. (MH/PRS/391/VOL.2/788)

### Laboratory Methods

Following demographic characterization, laboratory investigations were conducted to evaluate platelet indices and fibrinolytic markers across study groups. Standardized protocols were employed to ensure reproducibility and accuracy. Platelet indices (PLT, MPV, PDW, PCT) were determined using the Sysmex XP-300 haematology analyser (Obebinaru *et al*., 2023). Fibrinolytic markers, including D-dimer and fibrinogen, were quantified using Sandwich ELISA (Stang, 2013). Malaria diagnosis was confirmed by thick blood film microscopy with Giemsa staining (Torres *et al*., 2018), while COVID-19 status was verified by RT-PCR from nasopharyngeal swabs (Tsujimoto *et al*., 2021). These laboratory procedures provided the basis for subsequent statistical analyses of sex-specific hemostatic responses.

### Statistical Analysis

Data were analysed using SAS 9.4. Descriptive statistics (mean ± SD), one-way ANOVA with Tukey’s HSD post-hoc test, independent t-tests for sex comparisons, and Pearson correlation coefficients were performed. A p-value < 0.05 was considered statistically significant. Graphical representations (box plots and density curves) were generated using JMP software.

## Results

### Demographic Characteristics

A total of 220 participants were recruited in Port Harcourt, Nigeria, between November 2021 and March 2022, and stratified into four groups: healthy controls (n=55), malaria-positive (n=55), COVID-19-positive (n=55), and COVID-19+malaria co-infected (n=55). Each group was balanced by sex (22 females and 33 males), with a mean age of approximately 42 years (range: 20–65). Table 1 summarizes the demographic distribution across study groups, confirming comparability in age and sex composition.

**Table 1:** Demographic Characteristics.

| **Parameters** | **Controls**<br>**(n=55)** | **Malaria (n=55)** | **COVID-19**<br>**(n=55)** | **Co-infection**<br>**(n=55)** |
| --- | --- | --- | --- | --- |
| **Females** | 22 | 22 | 22 | 22 |
| **Males** | 33 | 33 | 33 | 33 |
| **Age**<br>**(mean ± SD, yrs)** | 41.8 ± 10.9 | 42.5 ± 11.2 | 42.0 ± 11.5 | 42.2 ± 11.6 |
| **Age Range (yrs)** | 20–65 | 20–65 | 20–65 | 20–65 |
| **Recruitment**<br>**Location** | Port Harcourt | Port Harcourt | Port Harcourt | Port Harcourt |
| **Recruitment**<br>**Period** | Nov 2021–Mar<br>2022 | Nov 2021–Mar<br>2022 | Nov 2021–Mar<br>2022 | Nov 2021–Mar<br>2022 |

**Table 2:**
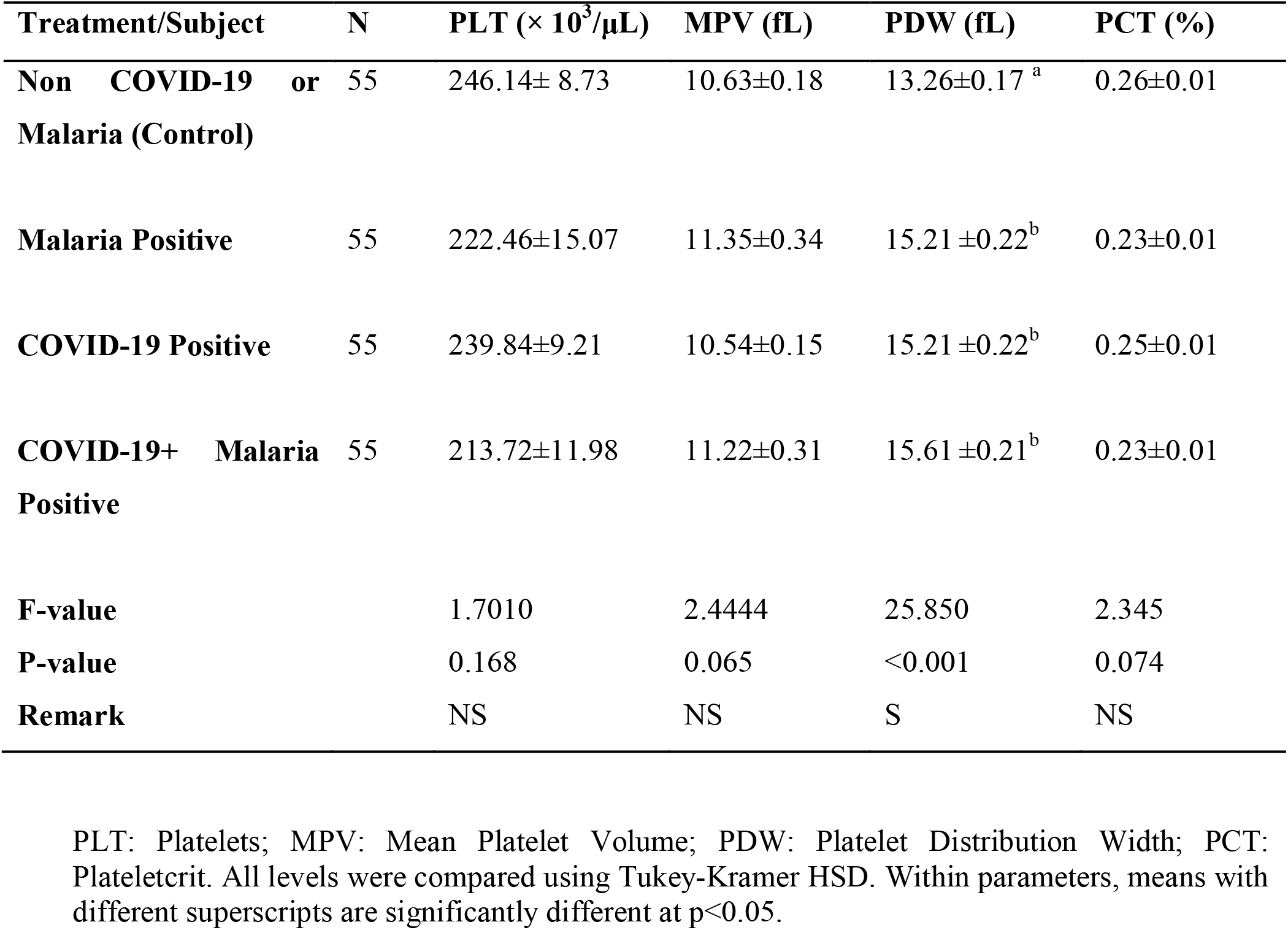
Mean ± SD of Platelet Indices of Study Subjects Infected with COVID-19 and Malaria. PDW was significantly higher in all infected groups compared to controls (p < 0.001), with the highest value in the co-infected group. No significant differences were observed in platelet count, MPV, or PCT across groups.

| Treatment/Subject | N | PLT ($\times 10^3/\mu\text{L}$) | MPV (fL) | PDW (fL) | PCT (%) |
| --- | --- | --- | --- | --- | --- |
| **Non COVID-19 or Malaria (Control)** | 55 | 246.14± 8.73 | 10.63±0.18 | 13.26±0.17<sup>a</sup> | 0.26±0.01 |
| **Malaria Positive** | 55 | 222.46±15.07 | 11.35±0.34 | 15.21 ±0.22<sup>b</sup> | 0.23±0.01 |
| **COVID-19 Positive** | 55 | 239.84±9.21 | 10.54±0.15 | 15.21 ±0.22<sup>b</sup> | 0.25±0.01 |
| **COVID-19+ Malaria Positive** | 55 | 213.72±11.98 | 11.22±0.31 | 15.61 ±0.21<sup>b</sup> | 0.23±0.01 |
| **F-value** |  | 1.7010 | 2.4444 | 25.850 | 2.345 |
| **P-value** |  | 0.168 | 0.065 | <0.001 | 0.074 |
| **Remark** |  | NS | NS | S | NS |

PLT: Platelets; MPV: Mean Platelet Volume; PDW: Platelet Distribution Width; PCT: Plateletcrit. All levels were compared using Tukey-Kramer HSD. Within parameters, means with different superscripts are significantly different at $p < 0.05$ .

**Table 3:** Mean ± SD of Some Fibrinolytic Markers of Study Subjects Infected with COVID-19 and Malaria. D-dimer levels were significantly elevated in the co-infected group compared to others (p = 0.040). Fibrinogen levels showed no overall significant difference among groups.

| **Treatment/Subject** | **N** | **Fibrinogen (ng/ml)** | **D-dimer (ng/ml)** |
| --- | --- | --- | --- |
| **Non COVID-19 or Malaria (Control)** | 55 | 55.36±2.03 | 319.86±51.93<sup>a</sup> |
| **Malaria Positive** | 55 | 54.11±1.56 | 429.60 ±59.87<sup>a</sup> |
| **COVID-19 Positive** | 55 | 57.48±2.57 | 431.34 ±55.51<sup>a</sup> |
| **COVID-19+ Malaria Positive** | 55 | 58.32±2.79 | 553.42 ± 59.74<sup>b</sup> |
| **F-value** |  | 0.7116 | 2.816 |
| **P-value** |  | 0.5461 | 0.040 |
| **Remark** |  | NS | S |

All levels were compared using Tukey-Kramer HSD. Within parameters, means with different superscripts are significantly different at $p < 0.05$ .

**Table 4:**
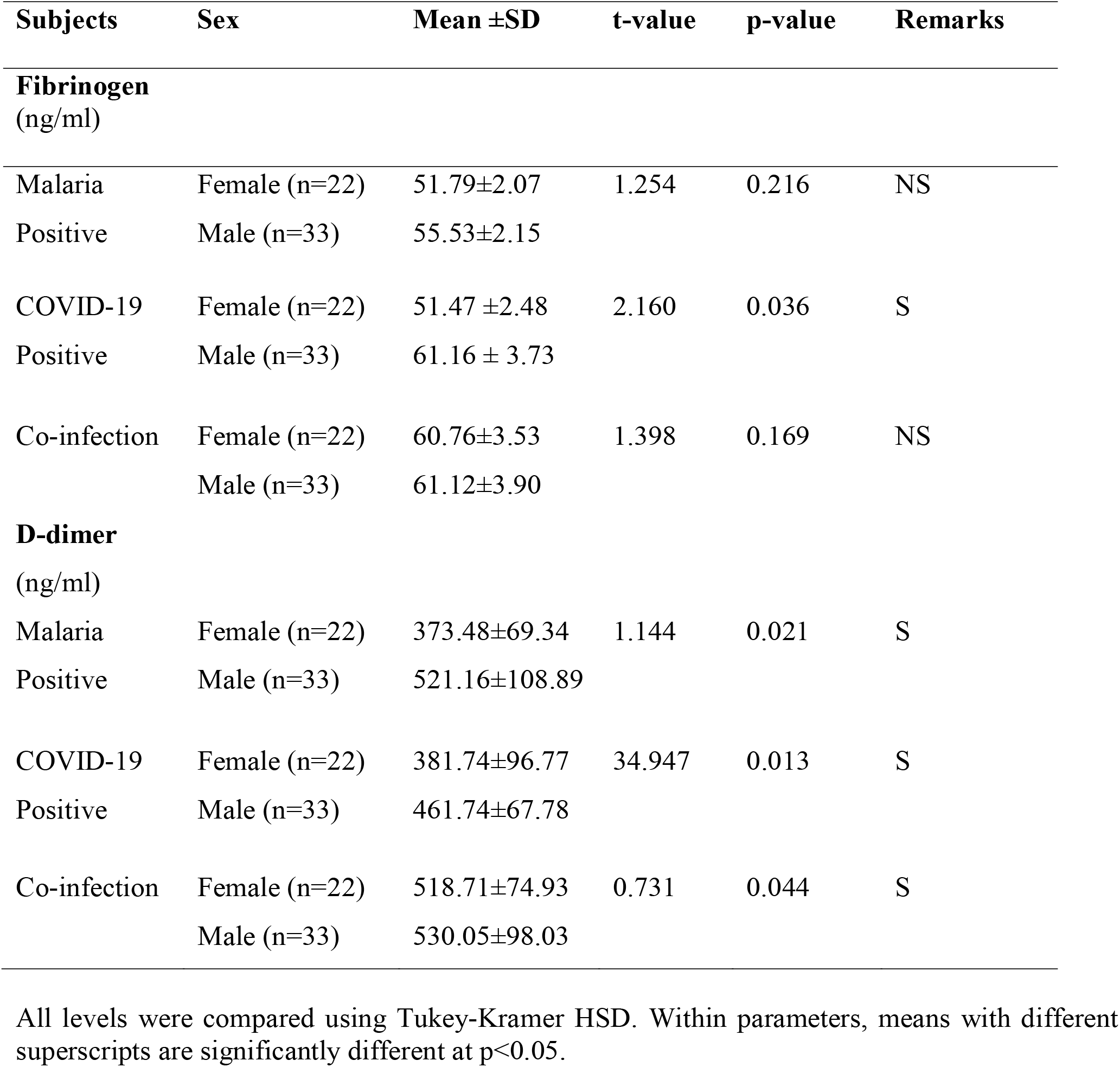
Values of Some Fibrinolytic Markers of Study Subjects Infected with COVID-19 and Malaria by Sex. Males demonstrated significantly higher D-dimer levels across all infected groups (p < 0.05). In COVID-19-positive subjects, males also had significantly higher fibrinogen levels (61.16 ± 3.73 ng/ml vs. 51.47 ± 2.48 ng/ml in females, p = 0.036). Platelet indices showed no significant sex differences.

| Subjects | Sex | Mean ±SD | t-value | p-value | Remarks |
| --- | --- | --- | --- | --- | --- |
| **Fibrinogen**<br>(ng/ml) |  |  |  |  |  |
| Malaria<br>Positive | Female (n=22) | 51.79±2.07 | 1.254 | 0.216 | NS |
| | Male (n=33) | 55.53±2.15 | | | |
| COVID-19<br>Positive | Female (n=22) | 51.47 ±2.48 | 2.160 | 0.036 | S |
| | Male (n=33) | 61.16 ± 3.73 | | | |
| Co-infection | Female (n=22) | 60.76±3.53 | 1.398 | 0.169 | NS |
| | Male (n=33) | 61.12±3.90 | | | |
| **D-dimer**<br>(ng/ml) |  |  |  |  |  |
| Malaria<br>Positive | Female (n=22) | 373.48±69.34 | 1.144 | 0.021 | S |
| | Male (n=33) | 521.16±108.89 | | | |
| COVID-19<br>Positive | Female (n=22) | 381.74±96.77 | 34.947 | 0.013 | S |
| | Male (n=33) | 461.74±67.78 | | | |
| Co-infection | Female (n=22) | 518.71±74.93 | 0.731 | 0.044 | S |
| | Male (n=33) | 530.05±98.03 | | | |

All levels were compared using Tukey-Kramer HSD. Within parameters, means with different superscripts are significantly different at $p < 0.05$ .

**Figure 1:**
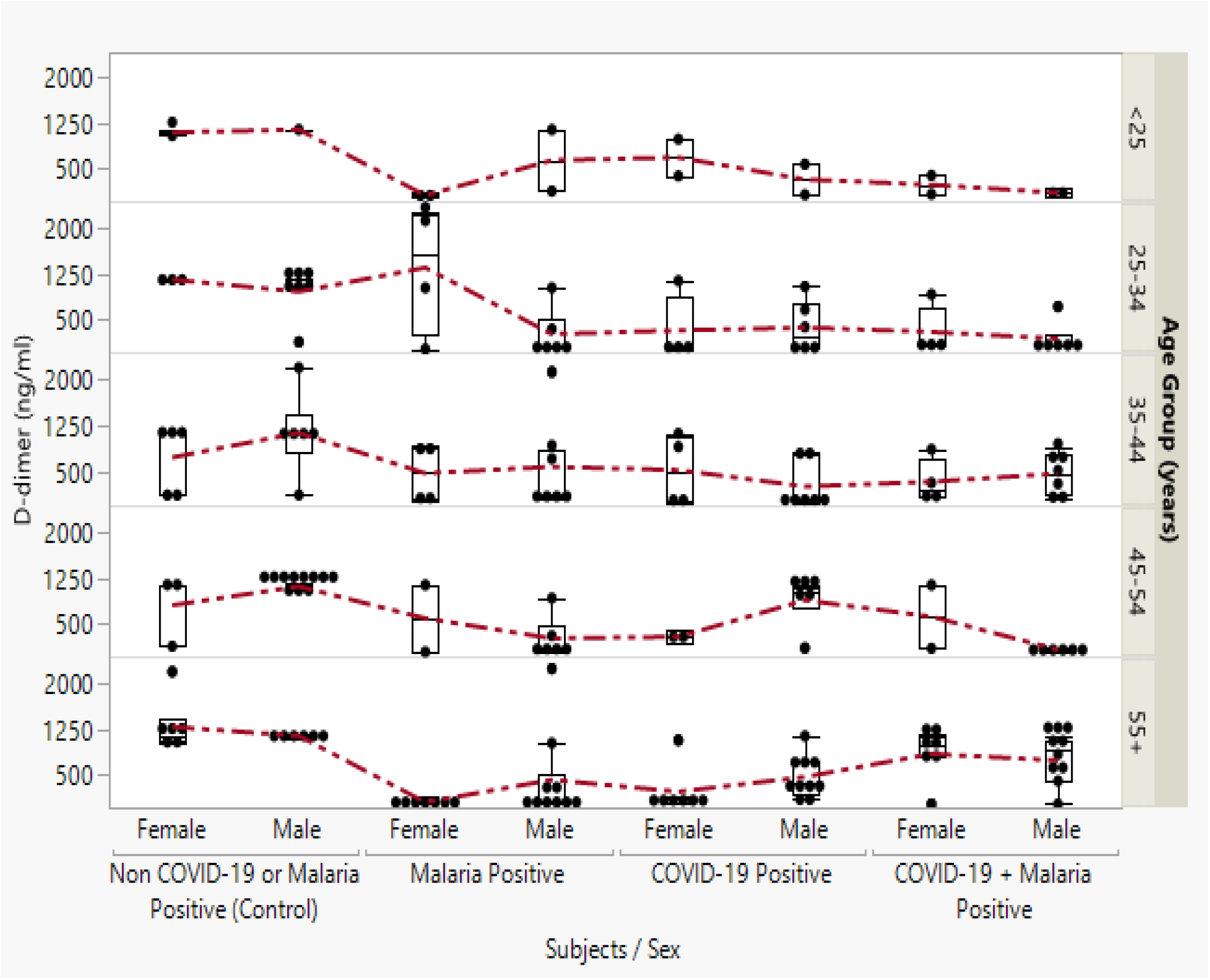
Box Plot of D-dimer in COVID-19 and Malaria Positive Subjects by Sex and Age Group.

**Table 5:** Correlation between Platelet Indices and Some Fibrinolytic Markers in the Study Groups. Strong positive correlation existed between PCT and PLT, and negative correlation between MPV and PLT across groups. No significant correlations were found between platelet indices and fibrinolytic markers (D-dimer or fibrinogen). Sex modified the distribution patterns more than age.

| Variable | by Variable | Correlation | P-value | Remark |
| --- | --- | --- | --- | --- |
| **MPV (fL)** | PLT ($\times 10^3/\mu\text{L}$) | -0.406 | 0.0035 | S |
| **PDW (fL)** | PLT ($\times 10^3/\mu\text{L}$) | 0.025 | 0.8640 | NS |
| **PDW (fL)** | MPV (fL) | -0.091 | 0.5310 | NS |
| **PCT (%)** | PLT ($\times 10^3/\mu\text{L}$) | 0.910 | <.0001 | S |
| **PCT (%)** | MPV (fL) | -0.102 | 0.4796 | NS |
| **PCT (%)** | PDW (fL) | -0.053 | 0.7140 | NS |
| **Fibrinogen (ng/ml)** | PLT ($\times 10^3/\mu\text{L}$) | 0.020 | 0.8895 | NS |
| **Fibrinogen (ng/ml)** | MPV (fL) | -0.114 | 0.4297 | NS |
| **Fibrinogen (ng/ml)** | PDW (fL) | 0.121 | 0.4033 | NS |
| **Fibrinogen (ng/ml)** | PCT (%) | -0.012 | 0.9366 | NS |
| **D-dimer (ng/ml)** | PLT ($\times 10^3/\mu\text{L}$) | -0.055 | 0.7060 | NS |
| **D-dimer (ng/ml)** | MPV (fL) | -0.003 | 0.9815 | NS |
| **D-dimer (ng/ml)** | PDW (fL) | 0.107 | 0.4605 | NS |
| **D-dimer (ng/ml)** | PCT (%) | -0.064 | 0.6595 | NS |
| **D-dimer (ng/ml)** | FIBRINOGEN (ng/ml) | 0.099 | 0.4958 | NS |

Abbreviations: PLT: Platelets; MPV: Mean Platelet Volume; PDW: Platelet Distribution Width; PCT: Plateletcrit, Significance Levels: $p < 0.01$ ; $p < 0.0001$ .

**Figure 2:**
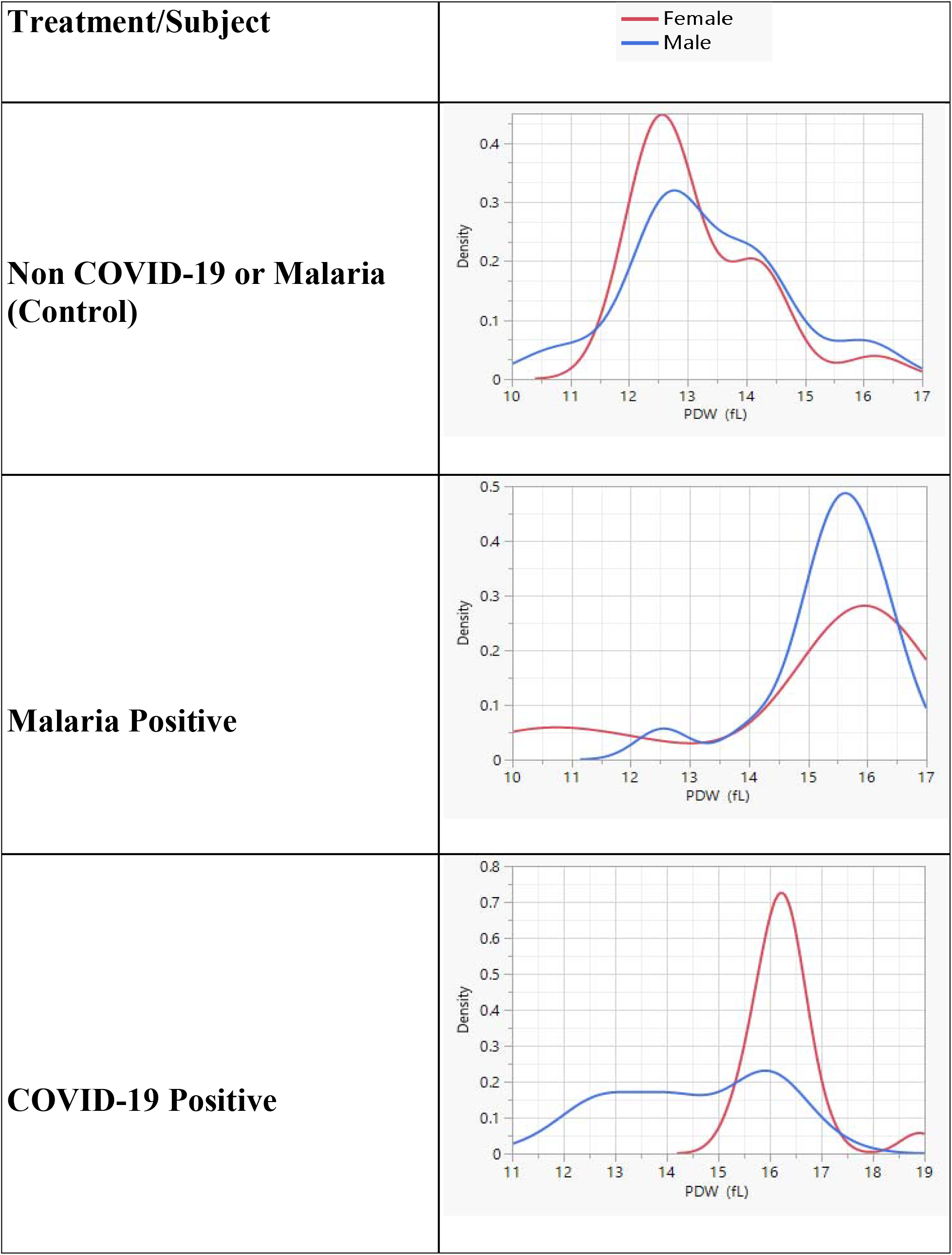
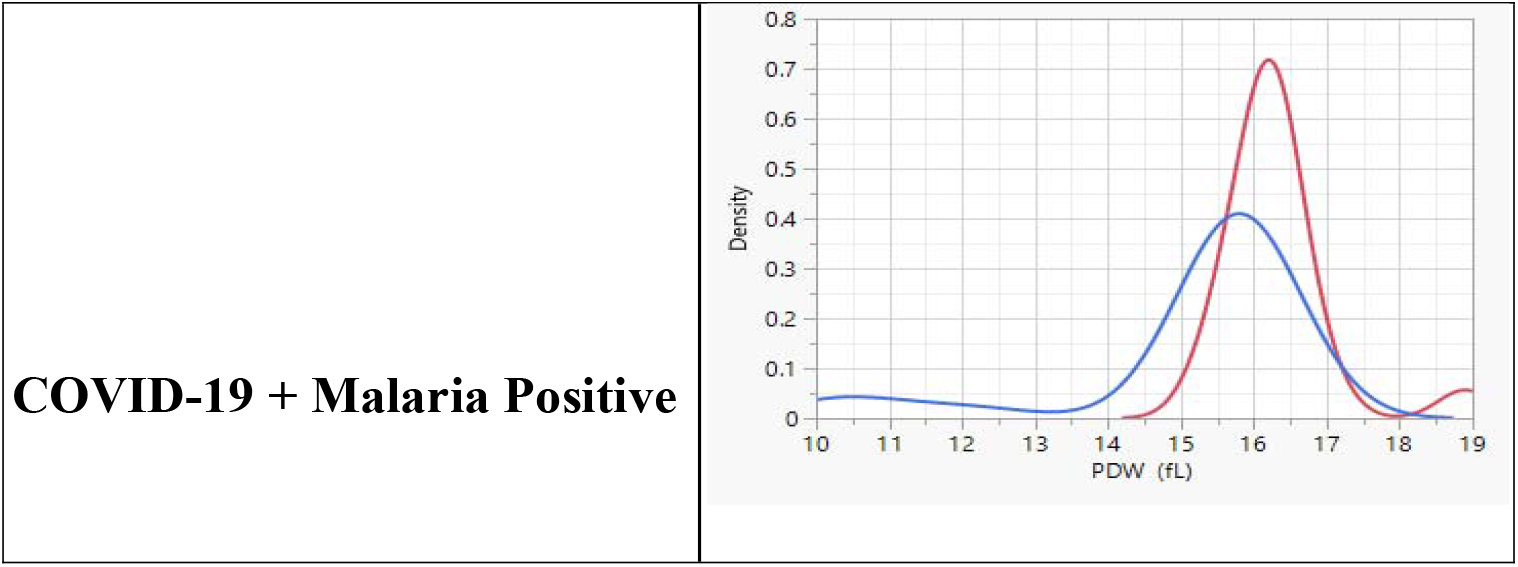
Comparison of Densities of Platelet Distribution Width (PDW) in COVID-19 and Malaria Positive Subjects by Sex.

**Figure 3:**
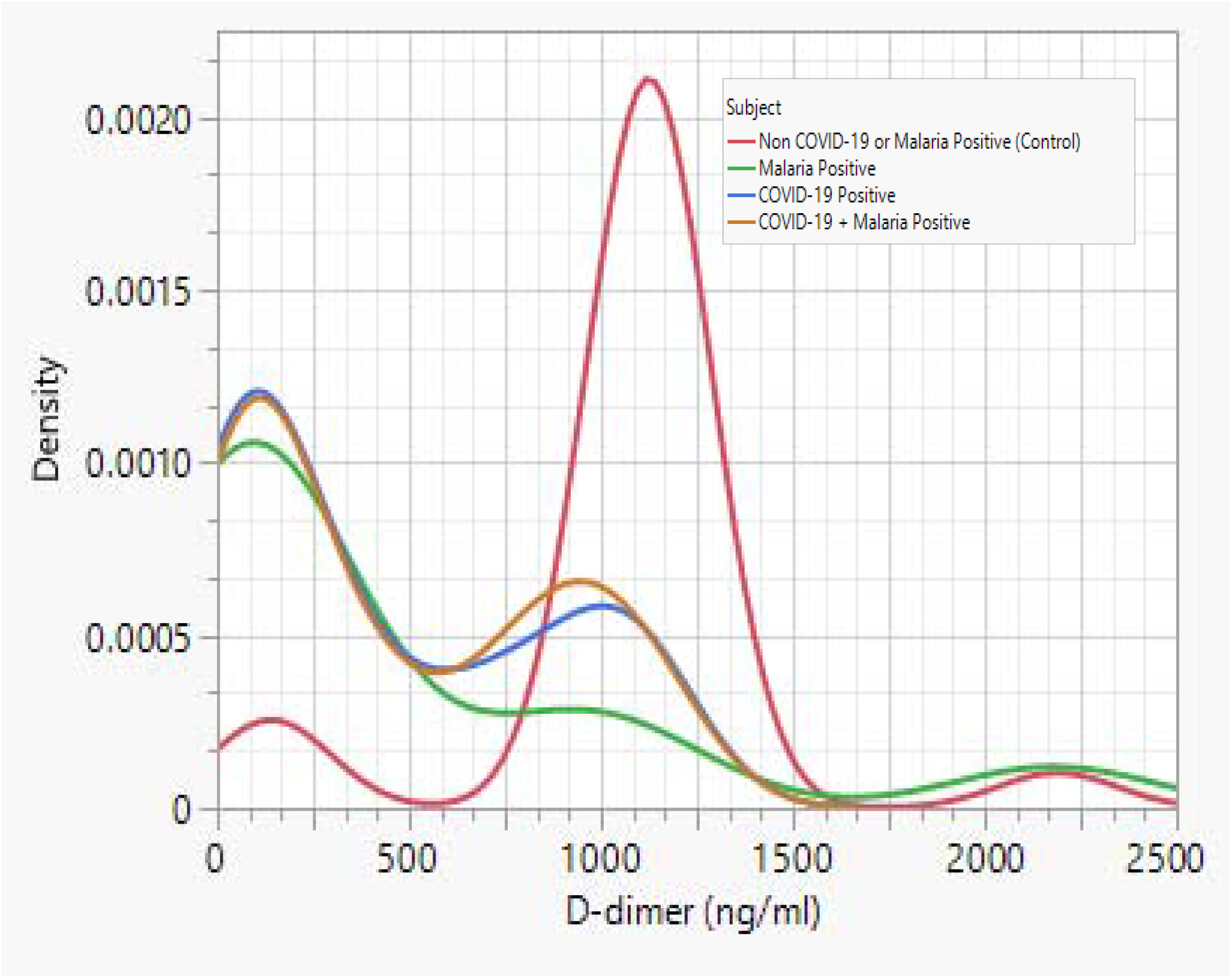
Comparison of Densities of D-dimer in COVID-19 and Malaria Positive Subjects.

## Discussion

This study reveals important sex-specific differences in hemostatic activation during mild COVID-19 and malaria infections. The consistent elevation of D-dimer in males across infected groups aligns with higher ACE2 expression and immunological differences between sexes. The pronounced increase in PDW across infected groups indicates significant platelet activation and turnover, even in mild/asymptomatic cases.

The highest D-dimer levels in co-infected subjects underscore the synergistic procoagulant effect of the two pathogens, consistent with endothelial damage and cytokine-mediated pathways described in the literature. PDW emerged as a robust marker of infection status, while its combination with D-dimer may enhance early identification of individuals at risk of coagulopathy in resource-constrained settings.

Strengths of this study include the well-matched case-control design in a real-world tropical co-endemic setting and comprehensive sex-stratified analysis. Limitations include the cross-sectional design and focus on mild cases, which may not fully represent severe disease dynamics.

## Conclusion

Males exhibit heightened fibrinolytic responses in COVID-19 and malaria co-infection. PDW and D-dimer are valuable, accessible biomarkers for screening and monitoring mild infections in tropical regions. Routine incorporation of these parameters in haematology panels could improve early risk stratification and clinical management in co-endemic areas.

### Recommendations

1. Integrate PDW and D-dimer into routine laboratory assessment for febrile patients in malaria/COVID-19 co-endemic zones.
2. Prioritise closer monitoring of male patients for coagulopathy.
3. Further longitudinal studies to evaluate predictive value for progression to severe disease.

## Data Availability

All data produced in the present work are contained right within the manuscript text, figures, and its supporting tables.

